# Gender-Specific Long-Term Prognostic Values of QRS Duration, QT Interval, and QTc from Automated ECG Analysis for Mortality and Adverse Outcomes in Patients Hospitalized for Heart Failure

**DOI:** 10.1101/2021.07.09.21260281

**Authors:** Jiandong Zhou, Sandeep S Hothi, Jeffery Shi Kai Chan, Sharen Lee, Wing Tak Wong, Keith Sai Kit Leung, Abraham Ka Chung Wai, Kamalan Jeevaratnam, Tong Liu, Gary Tse, Qingpeng Zhang

**Affiliations:** School of Data Science, City University of Hong Kong, Hong Kong, Hong Kong, China; New Cross Hospital, Wolverhampton, United Kingdom; Cardiovascular Analytics Group, Laboratory of Cardiovascular Physiology, Hong Kong, China; School of Life Sciences, The Chinese University of Hong Kong, Hong Kong, China; Emergency Medicine Unit, Li Ka Shing Faculty of Medicine, The University of Hong Kong, Hong Kong, China; Faculty of Health and Medical Sciences, University of Surrey, Guildford, United Kingdom; Tianjin Key Laboratory of Ionic-Molecular Function of Cardiovascular Disease, Department of Cardiology, Tianjin Institute of Cardiology, Second Hospital of Tianjin Medical University, Tianjin 300211, China

**Author notes:** Correspondence to: *Prof. Gary Tse PhD FRCP*, Tianjin Key Laboratory of Ionic-Molecular Function of Cardiovascular Disease, Department of Cardiology, Tianjin Institute of Cardiology, Second Hospital of Tianjin Medical University, Tianjin 300211, China, Faculty of Health and Medical Sciences, University of Surrey, GU2 7AL, Guildford, United Kingdom, *Prof. Qingpeng Zhang PhD*, School of Data Science, City University of Hong Kong, Hong Kong, China. equal contributions (Jiandong Zhou, Sandeep S Hothi).

**Keywords:** heart failure, electrocardiography, gender-specific outcomes, prognosis, automated ECG

## Abstract

**Background:** Gender-specific prognostic values of electrocardiographic (ECG) measurements in patients hospitalized for heart failure (HF) are lacking, which we hence investigated in this study.

**Methods:** Patients admitted to a single tertiary center for HF between 1 January 2010 and 31 December 2016 without atrial fibrillation and with at least one baseline ECG were included. Automated ECG measurements were performed. The primary outcomes were all-cause and cardiovascular (CAD) mortality, and the secondary outcomes were stroke, and ventricular arrhythmia and sudden cardiac death (VA/SCD). The prognostic values of the heart rate, PR segment, QRS duration, PT interval, QT interval, and QTc were assessed. Gender-specific optimal cutoffs of the above measurements were identified with the maximally selected rank statistics approach.

**Results:** In total, 2718 patients (median age 77 years; 1302 males) were included with a median follow-up of 4.8 years; the females were significantly older (p<0.0001). Females had higher rates of all-cause (p=0.04) and CAD mortality (p=0.02), while males had higher rates of VA/SCD (p=0.02). Higher heart rate, longer PT interval, wider QRS, and longer QT interval and QTc predicted all-cause mortality in males, while only shorter PR segment, longer QRS duration and QTc predicted the same in females. Longer QRS duration, QT interval, and QTc predicted CAD mortality in males, while longer PT interval, wider QRS and longer QTc predicted the same in females. ECG measurements also predicted the secondary outcomes to different extents depending on genders.

**Conclusions:** Selected ECG measurements have significant gender-specific prognostic value in patients admitted for heart failure.

## Introduction

Described as an emerging epidemic over 20 years ago, an estimated 64.3 million people lives with heart failure globally (1). As a heterogeneous clinical syndrome often presenting with signs and symptoms of fluid overload in acute exacerbations, heart failure is associated with varying degrees and progression of structural and electrophysiological alterations of the heart, and numerous previous studies have shown arrhythmogenic cardiac remodeling in heart failure (2, 3).

Heart failure admissions are common and costly (4), necessitating good prognostic markers for optimal risk stratification and early intervention for high-risk patients. Aside from blood tests and echocardiography, electrocardiogram (ECG) is one of the essential tools in the investigation and management of heart failure, in-patient and out-patient alike (5-7). In particular, ECG is readily available, inexpensive, and could be useful indicators of new onset morbidity and mortality in heart failure (HF) patients (7, 8).

However, gender-specific data of the prognostic value of ECG measurements are lacking. Gender differences in electrophysiological measurements are well known -- physiological data in healthy subjects have shown that QRS duration is shorter in females compared to males (9) . As such, the generalizability of non-gender-specific prognosticators cannot be assumed. As such, this study aimed to explore the gender-specific associations of other ECG measurements such as ventricular rate, PR segment, QRS duration, PT interval, QT interval, QTc and their dichotomized characteristics with the adverse outcomes of all-cause mortality, cardiovascular mortality, stroke, and ventricular arrhythmia / sudden cardiac death (VA/SCD) in heart failure patients.

## Methods

### Study design and population

This study was approved by The Joint Chinese University of Hong Kong - New Territories East Cluster Clinical Research Ethics Committee. This was a retrospective, territory-wide cohort study of hospitalized patients with ECG measurements between 1^st^ January 2000 and 31^st^ December 2019 from a single tertiary center in Hong Kong, China. The patients were identified from the Clinical Data Analysis and Reporting System (CDARS), a territory-wide database that centralizes patient information from 43 local hospitals and their associated ambulatory and outpatient facilities to establish comprehensive medical data, including clinical characteristics, disease diagnosis, laboratory results and drug treatment details. The system has been previously used by both our team and other teams in Hong Kong (10-12). Patients demographics, prior comorbidities, hospitalization characteristics before and after initial ECG measurement date, medication prescriptions, laboratory examinations of complete blood counts, biochemical renal and liver function tests, lipid and glucose tests were extracted. The *International Classification of Diseases, Ninth Revision, Clinical Modification* (ICD-9-CM) codes for comorbidities are detailed in the **Supplementary Table**

### Automated ECG measurements

Automatically measured parameters from ECG related to the P, Q, R, S and T-wave were extracted. The baseline ECG obtained on the first HF admission was selected.

### Outcomes and statistical analysis

The primary outcomes were all-cause mortality and cardiovascular mortality, and secondary outcomes include VA/SCD and stroke with follow-up until 31^st^ December 2019 (**Figure 1**). Mortality data were obtained from the Hong Kong Death Registry, a population-based official government registry with the registered death records of all Hong Kong citizens linked to CDARS. Cardiovascular mortality outcome was identified in death registry with *International Classification of Diseases, Tenth Revision, Clinical Modification* (ICD-10-CM) codes I10-I79. There was no adjudication of the outcomes as this relied on the ICD-9 coding or a record in the death registry. However, the coding was performed by the clinicians or administrative staff, who were not involved in the mode development. Patients without 12-leads ECG measurements and those with prior AF were excluded. The remaining cohort included 2718 patients.

**Figure 1.**
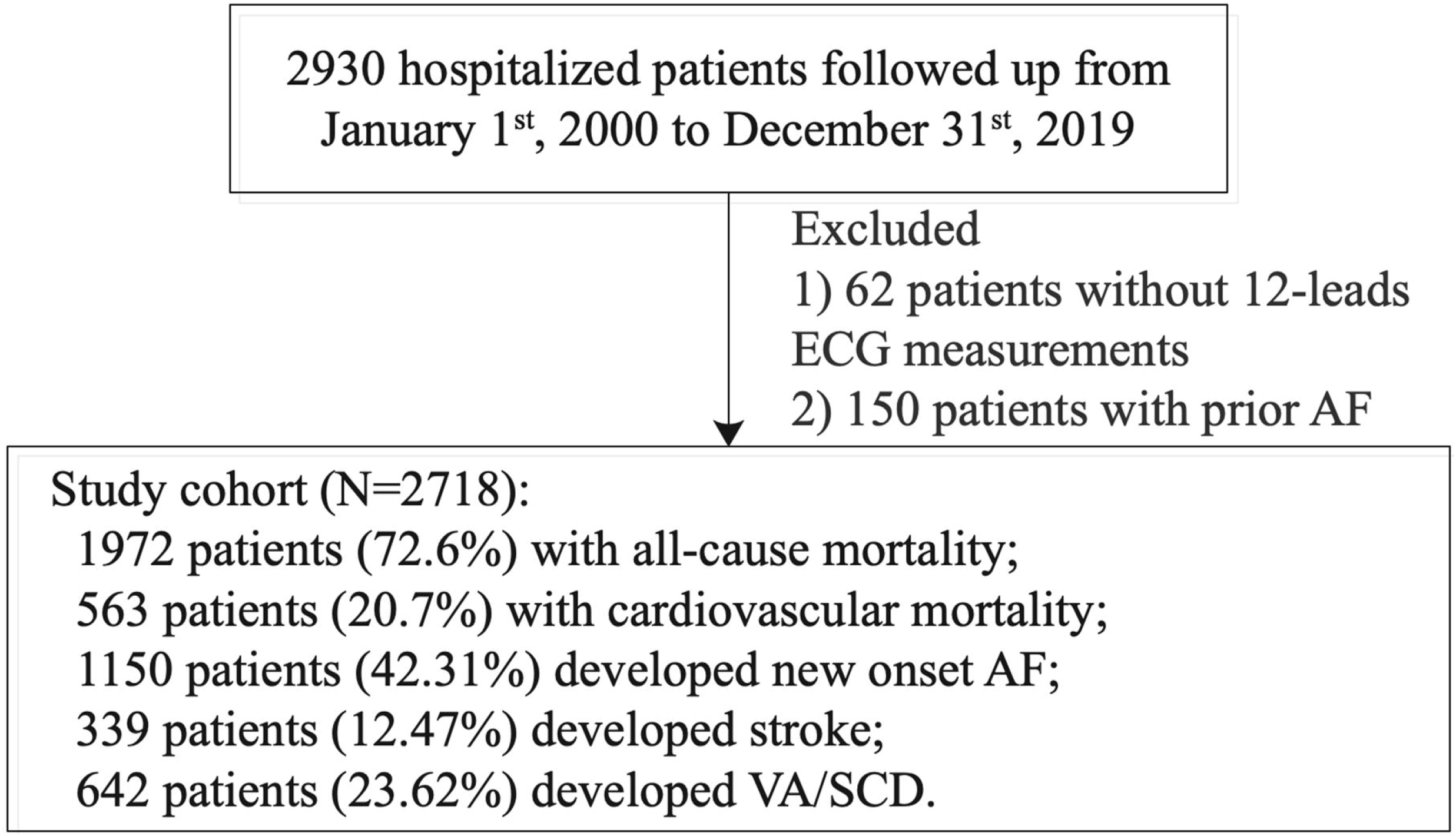
Study flowchart.

Continuous variables were presented as median (95% confidence interval [CI] or interquartile range [IQR]) and categorical variables were presented as count (%). The Mann-Whitney U test was used to compare continuous variables. The χ2 test with Yates’ correction was used for 2×2 contingency data. Univariate Cox regression models identify the significant risk factors of the primary and secondary outcomes. Hazard ratios (HRs) with corresponding 95% CIs and P-values were reported. There was no imputation performed for missing data. No blinding was performed for the predictor as the values were obtained from the electronic health records automatically. Gender-specific stratification performance of ventricular rate, PR segment, QRS duration, PT interval, QT interval, and QTc and their dichotomized characteristics (optimal cut-offs) for the primary and secondary outcomes were uncovered. The maximally selected rank statistics from the ‘maxstat’ R package is used to determine the optimal cut-off point for continuous variables (13). The maximally selected rank statistics approach is an outcome-oriented method providing a value of cutpoint that correspond to the most significant relation with outcome. Adjusted cubic spline model of the associations between ventricular rate, PR segment, QRS duration, PT interval, QT interval, and QTc and adverse risks of the primary and secondary outcomes were provided. All statistical tests were two-tailed and considered significant if p value<0.001. All statistical analyses were performed using RStudio software (Version: 1.1.456) and Python (Version: 3.6).

## Results

### Basic characteristics

Over a median follow-up of 4.8 (1.9-9.0) years, a total of 2718 patients with heart failure were enrolled (**Table 1**), of which 1302 (47.9%) were males. Female patients were significantly older, exhibited significantly higher CHA2DS2-VASc score (p<0.0001) and Charlson score (p<0.0001), albeit only with a very small absolute mean difference for the latter. More female patients suffered from hypertension and ischaemic heart disease, as well as using calcium channel blockers, statins and fibrates, and sodium-glucose cotransporter 2 inhibitors. The mean QRS duration was significantly longer in male patients.

**Table 1.** Gender-specific baseline and clinical characteristics of study population. * for p ≤ 0.05, ** for p ≤ 0.01, *** for p ≤ 0.001.

| Characteristics | All patients (N=2718)<br>Median (IQR);N or Count(%) | Males (N=1302)<br>Median (IQR);N or Count(%) | Females (N=1416)<br>Median (IQR);N or Count(%) | P value |
| --- | --- | --- | --- | --- |
| ***Outcomes*** | - | - | - | - |
| All-cause mortality | 1972(72.55%) | 932(71.58%) | 1040(73.44%) | 0.6859 |
| Cardiovascular mortality | 563(20.71%) | 250(19.20%) | 313(22.10%) | 0.1425 |
| Stroke | 339(12.47%) | 157(12.05%) | 182(12.85%) | 0.6206 |
| VA/SCD | 642(23.62%) | 339(26.03%) | 303(21.39%) | 0.0285* |
| ***Demographics*** |  |  |  |  |
| Baseline age, years | 77.36(66.94-84.3);n=2718 | 74.52(63.59-81.61);n=1302 | 80.01(71.16-86.6);n=1416 | <0.0001*** |
| CHA-DS-VASc Score | 4.0(3.0-5.0);n=2718 | 3.0(2.0-4.0);n=1302 | 5.0(4.0-6.0);n=1416 | <0.0001*** |
| Charlson score | 5.0(4.0-6.0);n=2718 | 5.0(3.0-6.0);n=1302 | 5.0(4.0-7.0);n=1416 | <0.0001*** |
| ***Past comorbidities*** |  |  |  |  |
| Diabetes without chronic complication | 790(29.06%) | 364(27.95%) | 426(30.08%) | 0.3872 |
| Diabetes with chronic complication | 256(9.41%) | 138(10.59%) | 118(8.33%) | 0.0764 |
| Renal diseases | 345(12.69%) | 161(12.36%) | 184(12.99%) | 0.7071 |
| Systemic embolism | 7(0.25%) | 3(0.23%) | 4(0.28%) | 0.9109 |
| Hypertension | 1253(46.10%) | 546(41.93%) | 707(49.92%) | 0.0122* |
| Myocardial infarction | 353(12.98%) | 220(16.89%) | 133(9.39%) | <0.0001*** |
| Chronic renal failure | 72(2.64%) | 35(2.68%) | 37(2.61%) | 0.9994 |
| Liver diseases | 15(0.55%) | 7(0.53%) | 8(0.56%) | 0.8701 |
| Ventricular tachycardia/fibrillation | 135(4.96%) | 72(5.52%) | 63(4.44%) | 0.2525 |
| Dementia and Alzheimer | 14(0.51%) | 4(0.30%) | 10(0.70%) | 0.2396 |
| COPD | 386(14.20%) | 206(15.82%) | 180(12.71%) | 0.0505 |
| IHD | 1012(37.23%) | 544(41.78%) | 468(33.05%) | 0.0017** |
| PVD | 66(2.42%) | 25(1.92%) | 41(2.89%) | 0.1372 |
| Stroke/TIA | 365(13.42%) | 155(11.90%) | 210(14.83%) | 0.0577 |
| Gastrointestinal bleeding | 336(12.36%) | 159(12.21%) | 177(12.50%) | 0.886 |
| Cancer | 162(5.96%) | 81(6.22%) | 81(5.72%) | 0.6613 |
| Obesity | 46(1.69%) | 17(1.30%) | 29(2.04%) | 0.1851 |
| ***Medications*** |  |  |  |  |
| ACEI | 1259(46.32%) | 635(48.77%) | 624(44.06%) | 0.1462 |
| ARB | 257(9.45%) | 109(8.37%) | 148(10.45%) | 0.1053 |
| Calcium channel blockers | 1232(45.32%) | 543(41.70%) | 689(48.65%) | 0.0278* |
| Beta blockers | 1334(49.08%) | 626(48.07%) | 708(50.00%) | 0.5814 |
| Diuretics | 1092(40.17%) | 502(38.55%) | 590(41.66%) | 0.2964 |
| Nitrates | 749(27.55%) | 368(28.26%) | 381(26.90%) | 0.579 |
| Statins and fibrates | 829(30.50%) | 447(34.33%) | 382(26.97%) | 0.0028** |
| Anticoagulants | 503(18.50%) | 258(19.81%) | 245(17.30%) | 0.1774 |
| Antiplatelets | 1117(41.09%) | 555(42.62%) | 562(39.68%) | 0.3327 |
| Sodium-glucose cotransporter 2 inhibitors | 44(1.61%) | 29(2.22%) | 15(1.05%) | 0.0265* |
| Dipeptidyl peptidase-4 inhibitors | 207(7.61%) | 112(8.60%) | 95(6.70%) | 0.099 |
| Alpha-glucosidase inhibitors | 69(2.53%) | 31(2.38%) | 38(2.68%) | 0.7139 |
| ***ECG measurements*** |  |  |  |  |
| Mean ventrate | 77.0(67.0-90.0);n=2717 | 77.0(67.0-90.0);n=1302 | 77.0(66.0-90.0);n=1415 | 0.7873 |
| Mean PT interval | 176.0(157.0-199.0);n=2717 | 176.0(158.0-199.0);n=1302 | 175.0(157.0-199.0);n=1415 | 0.6441 |
| Mean PR segment | 79.0(63.0-99.0);n=2717 | 79.0(64.0-99.0);n=1302 | 78.0(62.0-98.5);n=1415 | 0.3488 |
| Mean QRS duration | 91.0(82.0-108.0);n=2717 | 96.0(86.0-116.0);n=1302 | 88.0(80.0-101.0);n=1415 | <0.0001*** |
| Mean QT interval | 393.0(360.0-428.0);n=2717 | 391.0(360.0-424.0);n=1302 | 396.0(360.0-432.0);n=1415 | 0.0739 |
| Mean QTc | 444.0(420.0-472.0);n=2717 | 443.0(418.0-473.0);n=1302 | 446.0(423.0-472.0);n=1415 | 0.1035 |

ACEI, angiotensin converting enzyme inhibitor. ARB, angiotensinogen II receptor blocker. COPD, chronic obstructive pulmonary disease. ECG, electrocardiographic. IHD, ischaemic heart disease. IQR, interquartile range. PVD, peripheral vascular disease. TIA, transient ischaemic attack.

### Gender-specific analysis of outcomes

In total, the endpoint of all-cause mortality was met in 1972 (72.6%; 932 (71.6%) males) patients, stroke in 339 (12.5%; 157 (12.1%) males) patients, and VA/SCD in 642 (23.6%; 339 (26.0%) males) patients. Kaplan-Meier curves of stroke, VA/SCD, CAD mortality and all-cause mortality stratified by gender were presented in **Figure 2**. Males had lower adverse risk of CAD mortality (HR: 0.91, 95% CI: [0.10, 0.99], P value=0.0375) and all-cause mortality (HR: 0.82, 95% CI: [0.23, 0.96], P value=0.0161) than females, while males had higher adverse risk of VA/SCD (HR>1, P value<0.05). Adjusted cubic spline models of ventricular rate, PR segment, QRS duration, PT interval, QT interval, and QTc were presented to uncover their associations with the adverse risks of all-cause mortality (**Figure 3**), CAD mortality (**Figure 4**), stroke (**Figure 6**), and VA/SCD (**Figure 7**).

**Figure 2.**
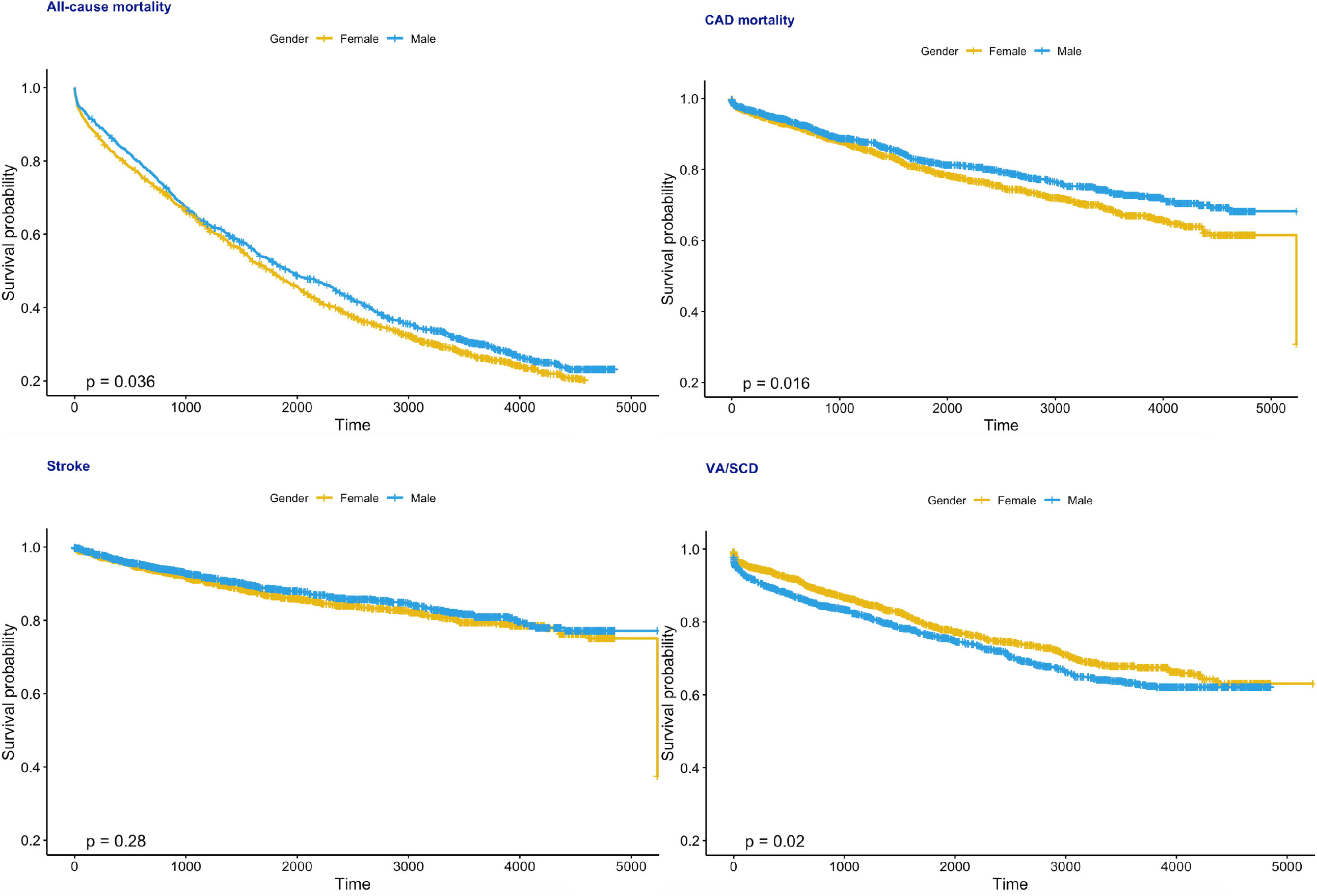
Kaplan-Meier curves of stroke, VA/SCD, CAD mortality and all-cause mortality stratified by gender.

**Figure 3.**
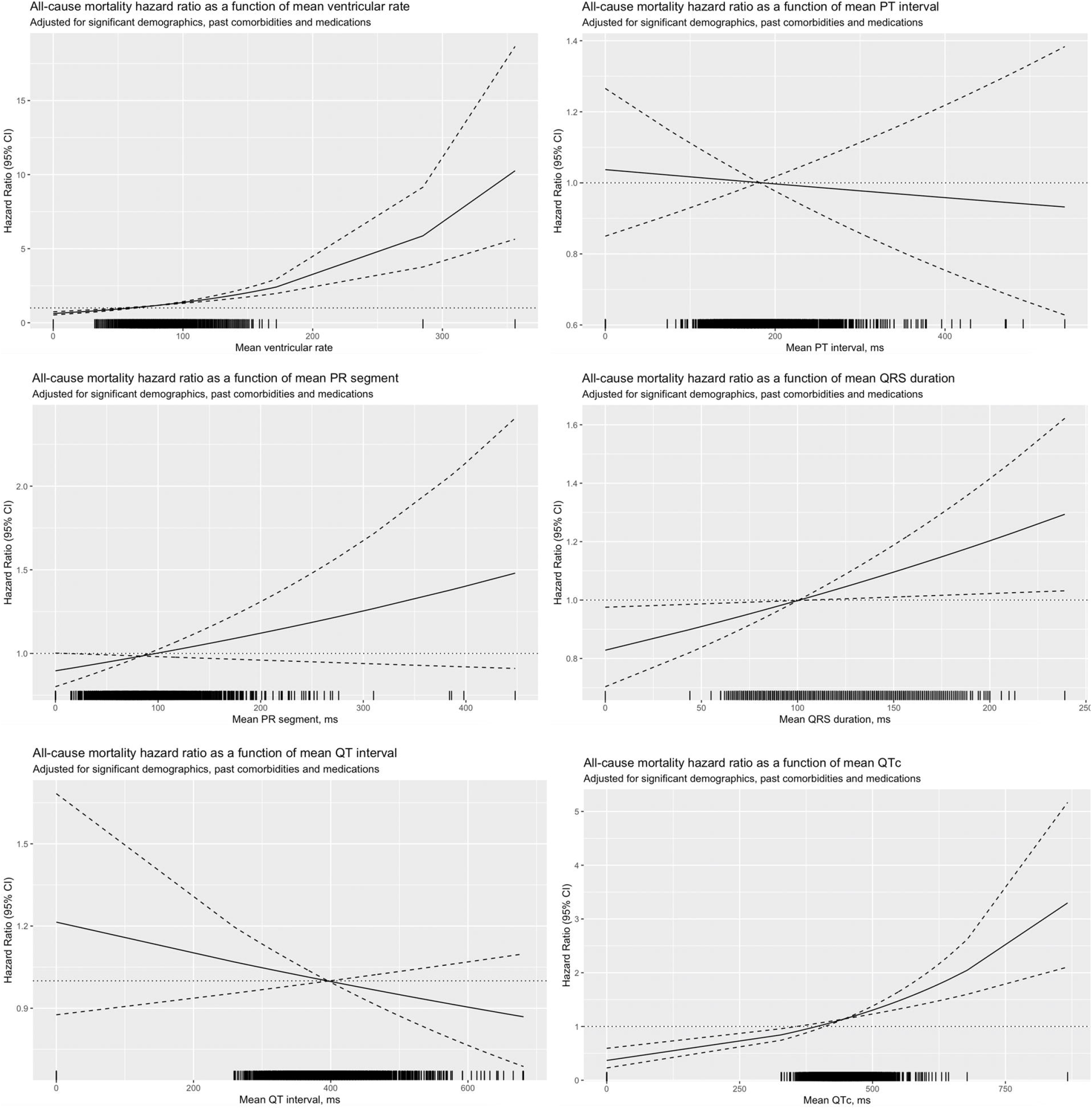
Adjusted cubic spline model of the associations between ventricular rate, PR segment, QRS duration, PT interval, QT interval, and QTc and adverse risks of all-cause mortality.

**Figure 4.**
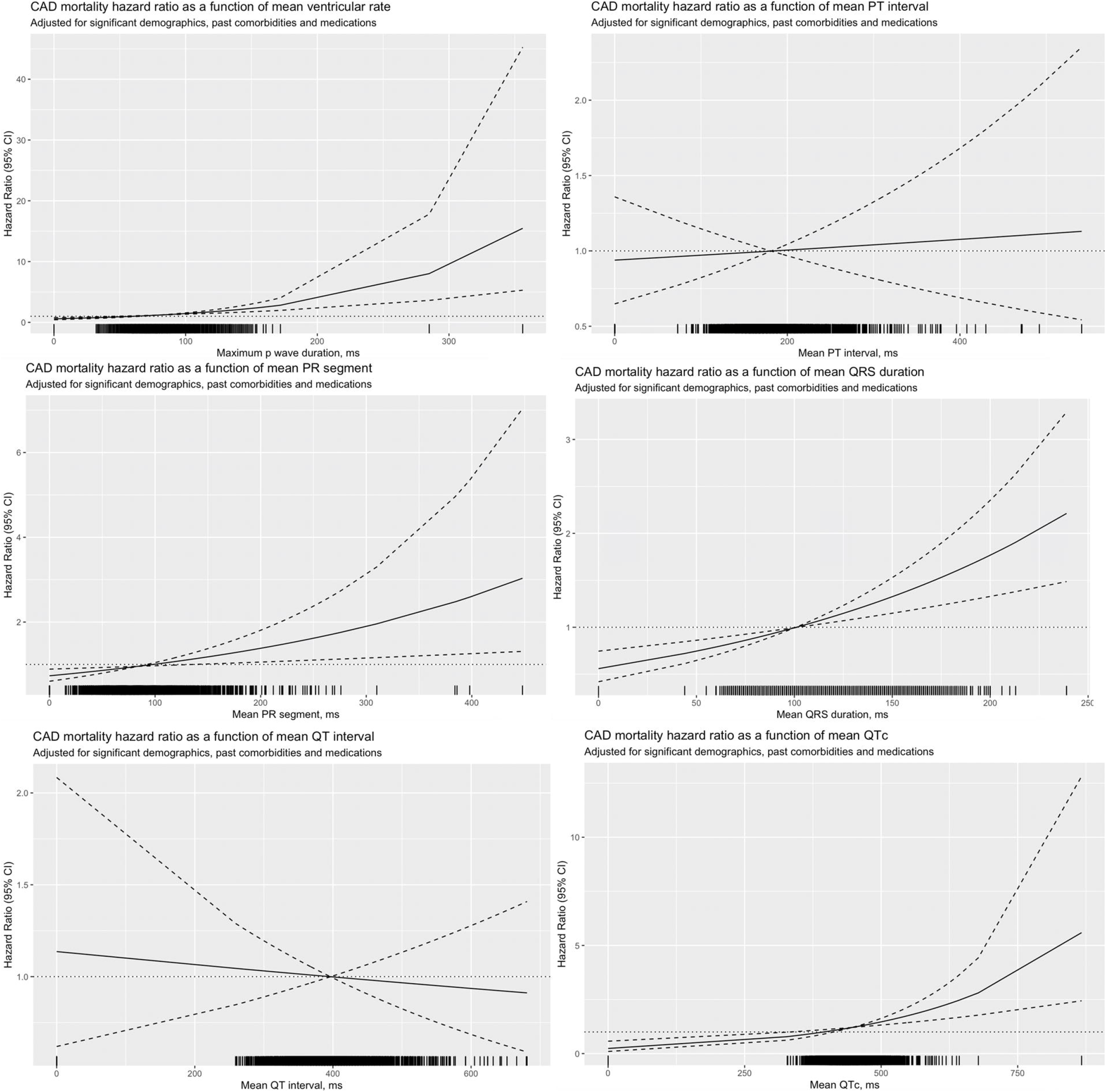
Adjusted cubic spline model of the associations between ventricular rate, PR segment, QRS duration, PT interval, QT interval, and QTc and adverse risks of CAD mortality.

**Figure 5.**
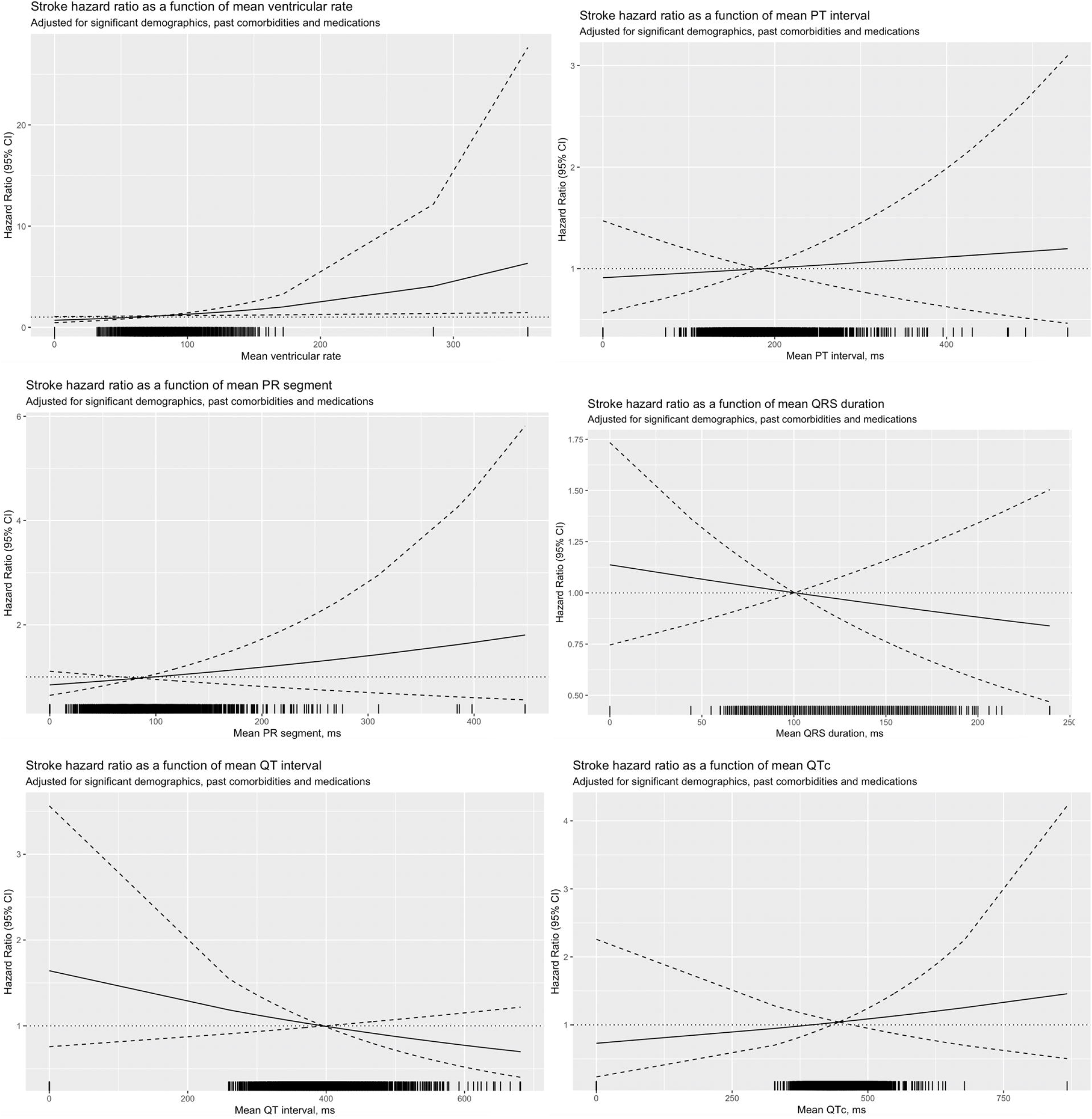
Adjusted cubic spline model of the associations between ventricular rate, PR segment, QRS duration, PT interval, QT interval, and QTc and adverse risks of stroke.

**Figure 6.**
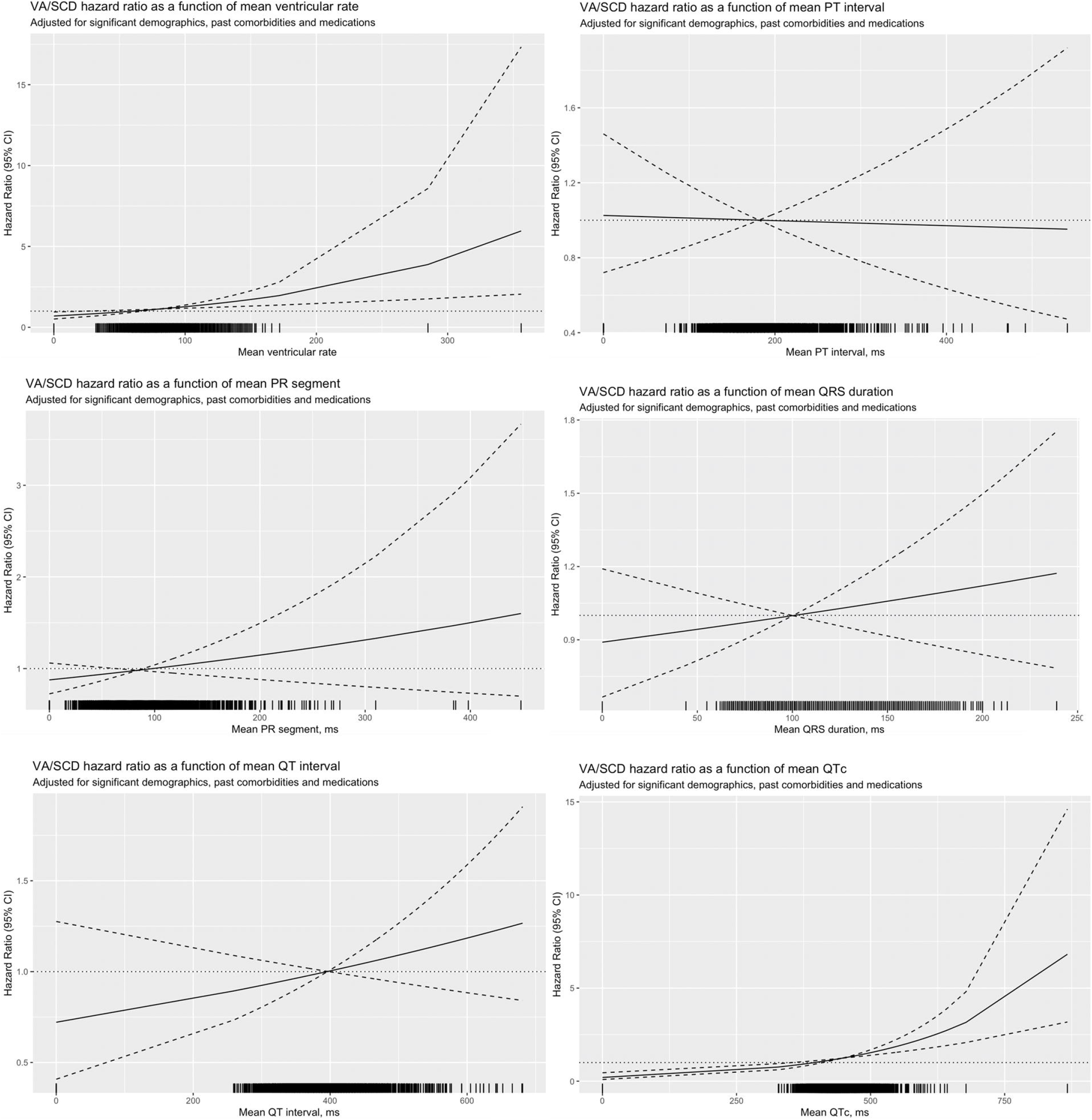
Adjusted cubic spline model of the associations between ventricular rate, PR segment, QRS duration, PT interval, QT interval, and QTc and adverse risks of VA/SCD.

### Gender-specific surface ECG markers of outcomes

Results of the gender-specific univariate Cox analysis are summarized in Supplementary Tables 2 and 3. A number of baseline comorbidities and medications were associated with the outcomes to varying extents. QTc length was predictive of all clinical outcomes except stroke in male patients. Higher heart rate and longer QTc were predictive of all-cause mortality in both genders, while longer PR segment, QRS duration and QTc were predictive of cardiovascular mortality in both genders. While longer PT interval and PR segment were predictive of all-cause mortality in males, they were not predictive in females. Similar discrepancy was observed in reverse for heart rate, which was predictive of cardiovascular mortality in females but not males.

**Table 2.** Gender-specific multivariate Cox analysis of the prediction strengths of dichotomized electrocardiographic markers. * for p ≤ 0.05, ** for p ≤ 0.01, *** for p ≤ 0.00

| **Males** |  |  |  |
| --- | --- | --- | --- |
| Dichotomized ECG measurements | All-cause mortality<br>HR [95% CI];P value | Dichotomized ECG measurements | Cardiovascular mortality<br>HR [95% CI];P value |
| Mean heart rate>95 BPM | 1.15[1.00-1.32];0.0455* | Mean heart rate>103 BPM | 1.14[0.87-1.50];0.3297 |
| Mean PT interval >187 ms | 1.18[1.03-1.35];0.0136* | Mean PT interval >185 ms | 1.28[1.00-1.65];0.0545 |
| Mean PR segment>107 ms | 1.04[0.90-1.19];0.6163 | Mean PR segment>89 ms | 1.21[0.92-1.59];0.1805 |
| Mean QRS duration>97 ms | 1.34[1.18-1.53];<0.0001*** | Mean QRS duration>101 ms | 1.59[1.24-2.05];0.0003*** |
| Mean QT interval>435 ms | 1.32[1.16-1.50];<0.0001*** | Mean QT interval>395 ms | 1.46[1.14-1.88];0.0029** |
| Mean QTc>428 ms | 1.52[1.33-1.74];<0.0001*** | Mean QTc>428 ms | 1.53[1.18-1.99];0.0012** |
| Dichotomized ECG measurements | Stroke<br>HR [95% CI];P value | Dichotomized ECG measurements | VA/SCD<br>HR [95% CI];P value |
| Mean heart rate>82 BPM | 1.00[0.70-1.41];0.9794 | Mean heart rate>93 BPM | 0.92[0.72-1.18];0.5263 |
| Mean PT interval >164 ms | 0.94[0.67-1.30];0.7009 | Mean PT interval >224 ms | 0.94[0.75-1.19];0.6281 |
| Mean PR segment>59 ms | 1.07[0.76-1.50];0.7156 | Mean PR segment>115 ms | 1.05[0.82-1.33];0.7145 |
| Mean QRS duration>90 ms | 0.75[0.54-1.03];0.0750 | Mean QRS duration>100 ms | 1.12[0.90-1.40];0.3171 |
| Mean QT interval>370 ms | 0.93[0.67-1.27];0.6367 | Mean QT interval>408 ms | 1.25[1.00-1.57];0.0468* |
| Mean QTc>484 ms | 0.79[0.54-1.15];0.2124 | Mean QTc>450 ms | 1.34[1.06-1.70];0.0149* |
| **Female** |  |  |  |
| Dichotomized ECG measurements | All-cause mortality<br>HR [95% CI];P value | Dichotomized ECG measurements | Cardiovascular mortality<br>HR [95% CI];P value |
| Mean heart rate>92 BPM | 1.13[0.99-1.28];0.0678 | Mean heart rate>106 BPM | 1.22[0.96-1.54];0.1084 |
| Mean PT interval >191 ms | 1.09[0.96-1.23];0.1766 | Mean PT interval >216 ms | 1.28[1.02-1.60];0.0296* |
| Mean PR segment>54 ms | 0.79[0.67-0.93];0.0053** | Mean PR segment>119 ms | 0.84[0.62-1.15];0.2785 |
| Mean QRS duration>87 ms | 1.17[1.03-1.32];0.0149* | Mean QRS duration>96 ms | 1.45[1.16-1.81];0.0011** |
| Mean QT interval>429 ms | 1.06[0.94-1.20];0.3206 | Mean QT interval>330 ms | 1.08[0.87-1.36];0.4801 |
| Mean QTc>443 ms | 1.40[1.24-1.58];<0.0001*** | Mean QTc>415 ms | 1.37[1.10-1.71];0.0057** |
| Dichotomized ECG measurements | Stroke<br>HR [95% CI];P value | Dichotomized ECG measurements | VA/SCD<br>HR [95% CI];P value |
| Mean heart rate>61 BPM | 1.13[0.83-1.54];0.4264 | Mean heart rate>83 BPM | 0.98[0.78-1.25];0.8981 |
| Mean PT interval >227 ms | 0.97[0.72-1.30];0.8388 | Mean PT interval >156 ms | 1.07[0.85-1.35];0.5606 |
| Mean PR segment>57 ms | 1.66[0.99-2.78];0.0527 | Mean PR segment>54 ms | 0.72[0.54-0.97];0.0329* |
| Mean QRS duration>98 ms | 1.24[0.93-1.67];0.1462 | Mean QRS duration>85 ms | 1.20[0.95-1.51];0.1226 |
| Mean QT interval>440 ms | 1.08[0.81-1.45];0.6023 | Mean QT interval>397 ms | 1.26[1.00-1.59];0.0525 |
| Mean QTc>443 ms | 1.47[1.10-1.97];0.0101* | Mean QTc>447 ms | 1.72[1.36-2.17];<0.0001*** |

1 CI, confidence interval. ECG, electrocardiographic. HR, hazard ratio. VA/SCD, ventricular arrhythmia and sudden cardiac death.

Multivariate gender-specific analysis of the prediction strengths of dichotomized ECGs for stroke, VA/SCD, and mortality risks in males and females were summarized in **Table 2**; all values were controlled for premorbid conditions and demographics that were associated with the outcomes. A wider QRS and longer QTc were both associated with higher all-cause and cardiovascular mortality, while a longer mean PT interval was predictive of all-cause mortality in males but cardiovascular mortality in females. Notably, a wider QRS was associated with the largest increase in risks of cardiovascular mortality amongst both genders, while a longer QTc was associated with the largest increase in risks of all-cause mortality. Interestingly, while a longer QTc was associated with drastically higher risk of stroke in female patients, no significant association was found in males. The important prognostic value of a long QTc was further reinforced by its association with VA/SCD in both genders.

## Discussion

In this territory-wide retrospective cohort study, we identified several ECG measurements which predicted adverse clinical outcomes in patients with heart failure. We also showed significant differences in clinical outcomes between genders.

Previous studies have found a variety of QRS measurements, such as P wave indices, QRS duration and QT intervals, to be predictive of mortality and adverse clinical outcomes in patients with heart failure (14-16). Elevated heart rate has also been shown to be predictive of mortality and adverse outcomes (17). These were corroborated by our findings. Mechanistically, anomalies of these indices represent disrupted cardiac depolarization, conduction and repolarization, in turn reflecting the severity of cardiac fibrosis and remodeling (3). Notably, though insignificant in males, QTc was predictive of stroke in women. While this might be explained as a reflection of the severity of heart failure and general condition, it is also possible that interactions between cerebral and autonomic dysfunctions, and electrophysiological and structural changes in heart failure may have played a role (18). Overall, with ECG being more easily accessible than blood markers and echocardiography in both in-patient and out-patient settings, our results reaffirm the value of 12-lead ECG in routine evaluation and prognostication of patients with heart failure. The current study also builds on recent efforts of exploring automated ECG measurements for prognostication in a variety of cardiovascular conditions (19, 20). Our study should serve as a good basis on which further investigations and wider application of automated ECG measurements may be based.

We showed important differences in clinical outcomes between sexes. Previous studies have demonstrated sex differences in clinical outcomes (21), and the reasons underlying such differences are multi-faceted, including genetic and aetiological differences, as well as different responses to pharmacotherapy (22-24). Importantly, analyses have demonstrated significant underrepresentation of women in clinical trials, which likely impacts the efficacy of guideline-driven medical therapy in female patients with heart failure (25). Additionally, findings regarding sex differences in mortality have been inconsistent. While the Framingham Heart Study and Olmsted County study, both carried out in the US, found significantly lower all-cause and cardiovascular mortality in female patients with heart failure respectively (26, 27),Sakata et al have shown the opposite in a Japanese cohort (28). Ceremers et al, on the other hand, have found neutral results in a Nordic cohort. Our results, having been derived from Asian patients, corroborated with the findings of Sakata et al. These pointed to a high likelihood of race-specific outcomes, as well as limited racial diversity and representation in heart failure trials. Particularly pertinent to the current study, this was supported by Mentz et al who showed, aside from variable representation of Asians in heart failure trials which limits their generalizability, considerable differences in disease profile, therapy, and outcomes among Asians as compared to their Caucasian counterparts (29). As such, further studies focusing on Asian patients with heart failure are warranted and optimize their treatments and prognostication specifically.

### Limitations

This study has several limitations. First, it is based on a territory-wide cohort from a single city in Hong Kong. This may limit the generalizability of our results to other populations. Second, due to the coding in CDARS and lack of echocardiographic data, the included cohort of patients with heart failure is heterogeneous, with a mix of different aetiologies and phenotypes. However, as a territory-wide study, our results closely reflect real-world practice and may still allow the general physician treating patients with heart failure to better assess them. Third, the accuracy of data recorded in CDARS cannot be assessed. Nonetheless, all the data was inputted by clinical staffs, and none of the authors were involved in data input. Data from CDARS have also been used in a number of peer-reviewed publications by our team and others. As such, we believe that CDARS remains a reasonably reliable source of territory-wide data.

## Conclusion

Outcomes of patients admitted for heart failure were significantly different between genders, and different ECG measurements significantly and independently predicted clinical outcomes in either gender.

## Data Availability

The data that support the findings of this study are available from the corresponding author upon reasonable request.

## Conflicts of Interest

None.

## Notes

### Competing Interest Statement

The authors have declared no competing interest.

### Funding Statement

No funding was obtained for this study

### Author Declarations

The Joint Chinese University of Hong Kong - New Territories East Cluster Clinical Research Ethics Committee

