## Supplementary Table for "Gender-Specific Long-Term Prognostic Values of QRS Duration, QT Interval, and QTc from Automated ECG Analysis for Mortality and Adverse Outcomes in Patients Hospitalized for Heart Failure"

**Supplementary Table 1. Codes for Comorbidities**

Diabetes without chronic complication 250	250.01	250.02	250.03	250.1	250.11	250.12	250.13	250.2	250.21	250.22	250.23	250.3	250.31	250.32	250.33	250.7	250.71	250.72	250.73

Diabetes with chronic complication	250.4	250.41	250.42	250.43	250.5	250.51	250.52	250.53	250.6	250.61	250.62	250.63

Renal diseases	582	582	582.1	582.2	582.4	582.8	582.81	582.89	582.9	583	583	583.1	583.2	583.4	583.6	583.7	585	585.1	585.2	585.3	585.4	585.5	585.6	585.9	586	588	588	588.1	588.8	588.81	588.89	588.9

Systemic embolism	444	444.01	444.09	444.1	444.2	444.21	444.22	444.8	444.81	444.89	444.9	445	445.01	445.02	445.8	445.81	445.89

Hypertension	401	401.1	401.9	402	402.01	402.1	402.11	402.9	402.91	403	403.01	403.1	403.11	403.9	403.91	404	404.01	404.02	404.03	404.1	404.11	404.12	404.13	404.9	404.91	404.92	404.93	405	405.01	405.09	405.1	405.11	405.19	405.9	405.91	405.99	437.2

Heart failure	428	428	428.1	428.2	428.2	428.21	428.22	428.23	428.3	428.3	428.31	428.32	428.33	428.4	428.4	428.41	428.42	428.43	428.9	398.91	402.01	402.11	402.91	404.01	404.03	404.11	404.13	404.91	404.93

Atrial fibrillation	427.31	429.4

Ventricular arrhythmias/sudden cardiac death 410	410.01	410.02	410.1	410.11	410.12	410.2	410.21	410.22	410.3	410.31	410.32	410.4	410.41	410.42	410.5	410.51	410.52	410.6	410.61	410.62	410.7	410.71	410.72	410.8	410.81	410.82	410.9	410.91	410.92	427.01	427.1	427.4	427.4	427.41	427.42	427.5	427.5	427.69	798	798.1	798.2

Liver diseases	456	456.1	456.2	572.2	572.3	572.4	572.8	571.4	571.5	571.6

Dementia and Alzheimer	331.82	290	290.1	290.11	290.12	290.13	290.2	290.21	290.3	290.4	290.41	290.42	290.43	290.8	290.9	294.2	294.1	294.11	294.21	332	46.1	333.4	340	42331	331.19	294.29

COPD	490	491	492	493	494	495	496	491.1	491.2	491.21	491.22	491.8	491.9	492.8	493.01	493.02	493.1	493.11	493.12	493.2	493.21	493.22	493.8	493.81	493.82	493.9	493.91	493.92	494.1	495.1	495.2	495.3	495.4	495.5	495.6	495.7	495.8	495.9

Peripheral vascular disease	250.7	443.9	443	443.1	443.2	443.21	443.22	443.23	443.24	443.29	443.8	443.81	443.82	443.89	441	443.9	785.4	V43.4

Stroke/TIA	Stroke/TIA	435	435.1	435.2	435.3	435.8	435.9	433.81	433.91	434	436	437	437.1	433.31	433.01	434.01	434.1	434.11	434.9	434.91	437.2	437.3	437.4	437.5	437.6	437.7	437.8	437.9	430	431	432	432.1	432.9

Gastrointestinal bleeding	531	531.2	531.4	531.6	532	532.2	532.4	532.6	533	533.2	533.4	533.6	534	534.2	534.4	534.6	535.01	535.11	535.21	535.31	535.41	535.51	535.61	535.71	562.02	562.03	562.12	562.13	569.3	569.85	569.86	578	578.1	578.9

IHD	410.01	410.02	410.1	410.11	410.12	410.2	410.21	410.22	410.3	410.31	410.32	410.4	410.41	410.42	410.5	410.51	410.52	410.6	410.61	410.62	410.7	410.71	410.72	410.8	410.81	410.82	410.9	410.91	410.92	411	411.1	411.8	411.81	411.89	413	413.1	413.9	414	414.01	414.02	414.03	414.04	414.05	414.06	414.07	414.1	414.11	414.12	414.19	414.2	414.3	414.4	414.8	414.9	410	412

Cancer 140-239

Obesity	278.01	278	278
